# The SweDen risk score- predicting death 1-year after myocardial infarction

**DOI:** 10.1101/2022.09.14.22279852

**Authors:** Rebecca T. Rylance, Philippe Wagner, Kevin K. W. Olesen, Jonas Carlson, Joakim Alfredsson, Tomas Jernberg, Margret Leosdottir, Pelle Johansson, Peter Vasko, Michael Maeng, Moman A. Mohammad, David Erlinge

**Author notes:** Correspondence: Rebecca T. Rylance, Department of Cardiology, Clinical Sciences, Lund University and Skåne University Hospital, Lund, Sweden, MSSC. The funding comes from the Swedish Research council and the Swedish HeartLung foundation.

## Abstract

**Objectives:** Our aim was to derive, based on the SWEDEHEART registry, and validate, using the Western Denmark Heart registry, a patient-oriented risk score, the SweDen score, which could calculate the risk of 1-year mortality following a myocardial infarction (MI).

**Methods:** The factors included in the SweDen score were age, sex, smoking, diabetes, heart failure, and statin use. These were chosen *a priori* by the SWEDEHEART steering group based on the premise that the factors were information known by the patients themselves. The score was evaluated using various statistical methods such as time-dependent receiver operating characteristics curves of the linear predictor, area under the curve metrics, Kaplan-Meier survivor curves, and the calibration slope.

**Results:** The area under the curve values were 0.81 in the derivation data and 0.76 in the validation data. The Kaplan-Meier curves showed similar patient profiles across datasets. The calibration slope was 1.03 (95% CI 0.99-1.08) in the validation data using the linear predictor from the derivation data.

**Conclusions:** The SweDen risk score is a novel tool created for patient use. The risk score calculator will be available online and presents mortality risk on a colour scale to simplify interpretation and to avoid exact life span expectancies. It provides a validated patient-oriented risk score predicting the risk of death at 1 year after suffering a MI, which visualizes the benefit of statin use and smoking cessation in a simple way.

## Introduction

Risk scores have been developed to aid in estimating the risk of new events or death after suffering a myocardial infarction (MI), motivate patients to adhere to treatment guidelines and lifestyle changes as well as optimize treatments for vulnerable patients.

The Global Registry of Acute Coronary Events (GRACE) score was based on 18 clusters in 14 countries gathering 10,000 acute coronary syndrome (ACS) patients yearly.^1^ In its first version, the GRACE score incorporated age, heart rate, systolic blood pressure, serum creatinine, Killip class, cardiac arrest at admission, deviations of the ST segment and cardiac enzyme levels to predict in-hospital mortality.^2^ The first TIMI score was developed for unstable angina/non-ST myocardial infarction to evaluate a composite end point of all-cause mortality, MI, and urgent revascularization.^3^ It consisted of 7 factors including age 65 years or older, having ≥ 3 coronary artery disease (CAD) risk factors such as hypertension, hypercholesterolemia, diabetes, family history of CAD or current smoker, prior coronary stenosis of 50% or more, prior ST-segment deviation on electrocardiogram at presentation, at least 2 angina events in the prior 24 hours, the use of aspirin in the prior 7 days, and elevated serum cardiac markers. However, these risk scores are not suitable for patients to use by themselves.

The Swedish Web-system for Enhancement and Development of Evidence-based care in Heart disease Evaluated According to Recommended Therapies (SWEDEHEART) registry started in 2009, and encompasses 95% of all acute first time or repeated MI cases in Sweden of those under the age of 80. Background characteristics such as age, body mass index, smoking status, electrocardiographic findings as well as other examinations, interventions, complications, discharge medications and diagnoses are prospectively collected. The Western Denmark Heart Registry contains similar information on patients.

In a world where patients seek knowledge and guidance online, we found the idea of a patient-oriented risk score both novel and intriguing. Therefore, the aim of this study was to develop a user-friendly risk score predicting death within 1 year after suffering a MI based on the Swedish and Danish populations.

## Methods

### Data selection

For this study, data from the 1st of January 2008 to the 27^th^ of May 2018 from the SWEDEHEART registry was selected, consisting of 247,904 MI cases. Patients who died during hospital stay or within 30 days after their MI were excluded. Patients with cancer or dementia, patients under the age of 55, and patients who received cardio-pulmonary resuscitation (CPR) on their way to hospital were excluded. For patients with current events, the last hospital stay per patient was selected, assuming that this represents the most valid patient information, and the final database consisted of 125,806 patients.

### Factors in model

The factors chosen for the SweDen score were chosen *a priori* by the SWEDEHEART steering group based on the premise that the factors should be clinically relevant information known by the patients themselves. These included age, sex, smoking (both current and previous), diabetes, heart failure, and being prescribed statins.

### Estimating the model

Age was treated as a continuous variable in the model. The categorical variables included in the model were categorized with a relevant reference group; if the patient had a condition that was associated with a higher risk, they were coded as ‘1’ and if they did not, they were coded as ‘0’. As such, having heart failure, diabetes, being male, being a current or previous smoker, or not being treated with a statin were associated with higher risks. A Cox model was fitted with the pre-selected factors to generate log coefficients. Log-minus-log survival plots and the Schoenfeld residuals were checked visually to ascertain model fit.

### Generating the risk score

The Framingham tutorial for clinical use was the basis for calculating the risk score.^4^ This involved several steps (Appendix). For the calculation of the points, the age variable was categorized into 5-year age groups and the midpoint in each age category was used. The youngest age group included people between 55 and 60 and therefore the midpoint for that age group was 57. The definition of a point was 5 years of aging, which was calculated by taking the log hazard coefficient for age produced by the model and multiplying it by 5 and hereby referred to as *B*. The number of points was calculated for each factor. The number of points for each increase into a higher age group was found by taking the difference between the midpoints in each age group minus the midpoint in the lowest age group, 57, and multiplying it by the log hazard coefficient for age and dividing it by *B*. For example, if a person were 78 years old, they would be in the age group from 75-79 and the midpoint in that group is 77. The number of points for being 78 years old was calculated to 4, (0.0780096*(77-57)/0.390048). The number of points for each categorical variable was produced similarly by taking the log hazard coefficient produced in the model for each particular variable and dividing by *B*. The diabetes variable produced a log hazard of 0.5153974, which constituted a 1-point increase if a person had diabetes (0.5153974*(1-0)/0.390048). The number was 1.32 and was rounded down to 1. Being male did not add an additional point however, 0.0894894*(1-0)/0.390048 was equal to 0.23 and was rounded down to zero. Being a previous smoker did not add an additional point either. The total points were summed, and each point total was associated with a risk.

### Validation

External validation involves a derivation dataset where the original analysis is performed and a validation dataset where the results are tested and verified.^5^ The risk score was derived with the SWEDEHEART registry data, and was therefore the derivation data. The Western Denmark Heart Registry, containing 45,003 patients with the same selection criteria as the SWEDEHEART registry was used to evaluate the SweDen risk score and was the validation dataset.^6^ Hazard ratios were produced for both populations.

Calibration and discrimination were two important concepts that were applied in the validation of the risk score model.^7^ Discrimination can be defined as the model’s ability to correctly separate low and high-risk patients.^8^ Patients who were predicted to be at a higher risk should have experienced higher event rates.

Time-dependent receiver operating characteristics (ROC) curves of the linear predictor, which are the weighted sum of the factors in the derivation and validation models were produced as well as area under the curve (AUC) metrics to evaluate discrimination. The AUC calculates the probability that in a pair of patients selected at random, the patient with the shorter survival time has the higher risk.^9^ The linear predictors were also plotted in histograms to visualize their spread.

Kaplan-Meier curves of the risk groups were graphed as an informal assessment of discrimination.^8^ The more widely separated the curves, the better the discrimination. The linear predictor was divided into quantiles at the 16^th^, 50^th^, and 84^th^ centiles. The percentages of patients in each of these risk groups, which can be thought of as good, fairly good, fairly poor, and poor risk groups, were compared between the derivation and validation data. Hazard ratios of these risk groups and their confidence intervals were also computed. Calibration may be defined in terms of prediction accuracy, i.e., how closely the survival in the validation data was captured by the model’s predictions from the derivation data.^8^ The calibration slope in the validation dataset was calculated by taking the coefficients produced from the derivation data and performing a Cox regression with them using the Danish data.

## Results

The results are presented for both derivation and validation datasets. Baseline demographics are presented in Table 1. Time-dependent ROC curves are presented (Figure 1) with AUC values of 0.81 in the derivation dataset and 0.76 in the validation dataset. Histograms visually demonstrate the spread of the centered linear predictor for the risk groups. No obvious outliers or irregularities were noted (Figure 3 - Appendix). Hazard ratios are presented in Table 2. All covariates were significant in the models except for previous vs. non-smoker, with a HR of 1.04 (95% CI 0.99-1.08) in the derivation data and an HR of 1.03 (95% CI 0.93-1.13) in the validation dataset. The Kaplan-Meier survival curves of the risk groups are depicted for both datasets (Figure 2). The percentages of patients in the 4 groups, representing good, fairly good, fairly poor, and poor risk of survival were 15.5%, 32.6%, 33.4% and 18.6% in the derivation dataset, and 15.5%, 34.5%, 34.0% and 16.0% in the validation dataset, respectively, showing similiar distributions of patient profiles in both datasets. The calibration slope was 1.03 (95% CI 0.99 1.08). The risk score resulted in a patient-based online calculator where an increasing number of points signifies an increasing risk of death (Figure 4). A total of 0 points means a very low risk of death whereas a total of 14 points conveys a very high risk of death. The risk of dying with 0 points was 1.8%, 1 point 2.6%, 2 points 3.8%, 3 points 5.6%, 4 points 8.1%, 5 points 11.8%, 6 points 16.9%, 7 points 23.9%, 8 points 33.2%, 9 points 44.9%, 10 points 58.5%, 11 points 72.7%, 12 points 85.3%, 13 points 94.1%, and 14 points 98.5%. The score can be accessed here: www.sweden-score.info/english.

**Table 1.** Baseline demographics SWEDEHEART registry (Derivation dataset) & Western Denmark registry (Validation dataset)

| Baseline | Derivation<br>n = 125806 | Validation<br>n = 45003 |
| --- | --- | --- |
| Male sex, n (%) | 80136 (63.7) | 30458 (67.7) |
| Age, mean (SD) | 73.3 ± 10.0 | 69.3 ± 8.7 |
| Diabetes, n (%) | 27874 (22.2) | 7833 (17.4) |
| Body Mass Index (kg/m<sup>2</sup>), mean (SD) | 26.9 ± 5.7 | 26.9 ± 4.6 |
| Non-smoker, n (%) | 59336 (47.2) | 15051 (33.4) |
| Previous smoker, n (%) | 45166 (35.9) | 16642 (37.0) |
| Current smoker, n (%) | 21304 (16.9) | 13310 (29.6) |
| Previous PCI | 28002 (21.6) | 1003 (2.2) |
| Previous CABG | 10552 (8.1) | 281 (0.6) |
| Heart failure, n (%) | 16514 (13.1) | 10930 (24.3) |
| Hypertension, n (%) | 69034 (54.9) | 26237 (58.3) |
| No statins, n (%) | 16323 (13.0) | 6114 (13.6) |
| P-creatinine (umol/L), mean (SD) | 94.4 ± 57.2 | 101.9 ± 684.4 |
| Lipid-lowering treatment | 111669 (88.8) | 20887 (46.4) |
| Aspirin | 116549 (92.6) | 39329 (87.4) |
| Ace-inhibitor | 75935 (60.4) | 20516 (45.6) |
| Beta blocker | 112652 (89.5) | 35809 (79.6) |

**Table 2.**
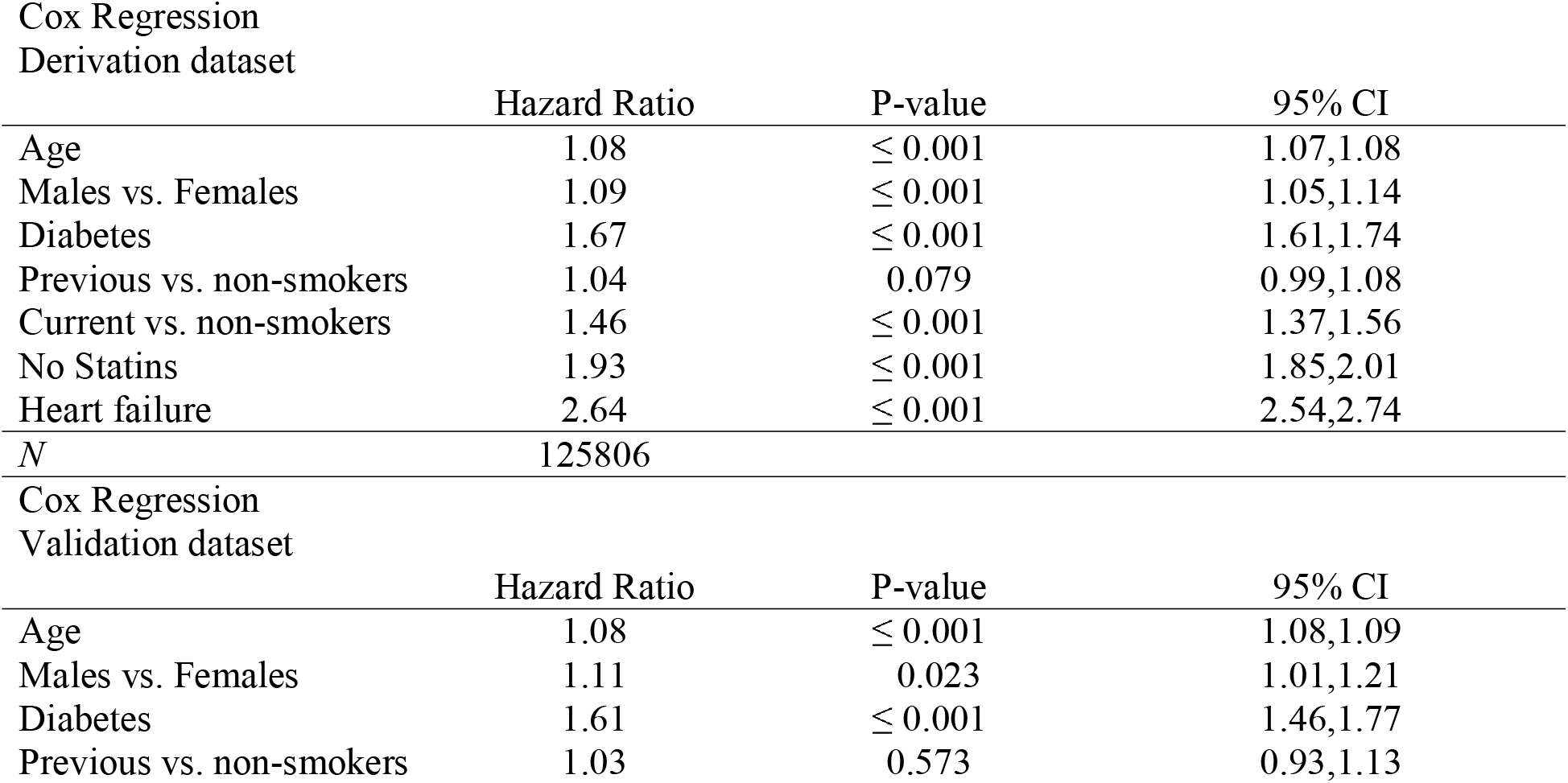
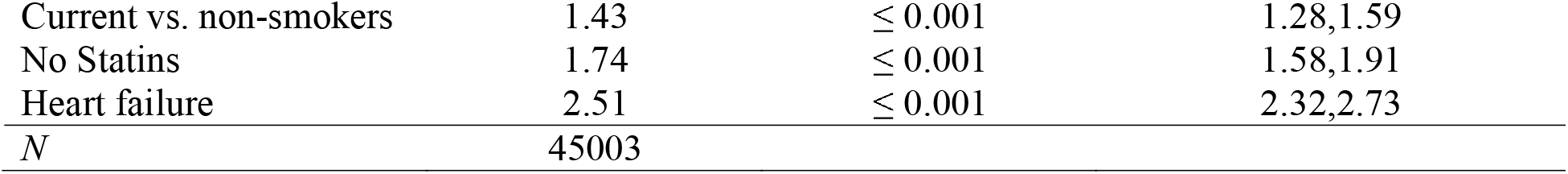
Hazard ratios from a model adjusted for age, gender, diabetes, smokers, statins and heart failure using Cox regression from the SWEDEHEART registry (Derivation dataset), and the Western Denmark registry (Validation dataset)

**Table 3.** Hazard ratios between the categorized linear predictor in risk groups from the SWEDEHEART registry (Derivation dataset) and Western Denmark registry (Validation dataset) Appendix

**Figure 1.**
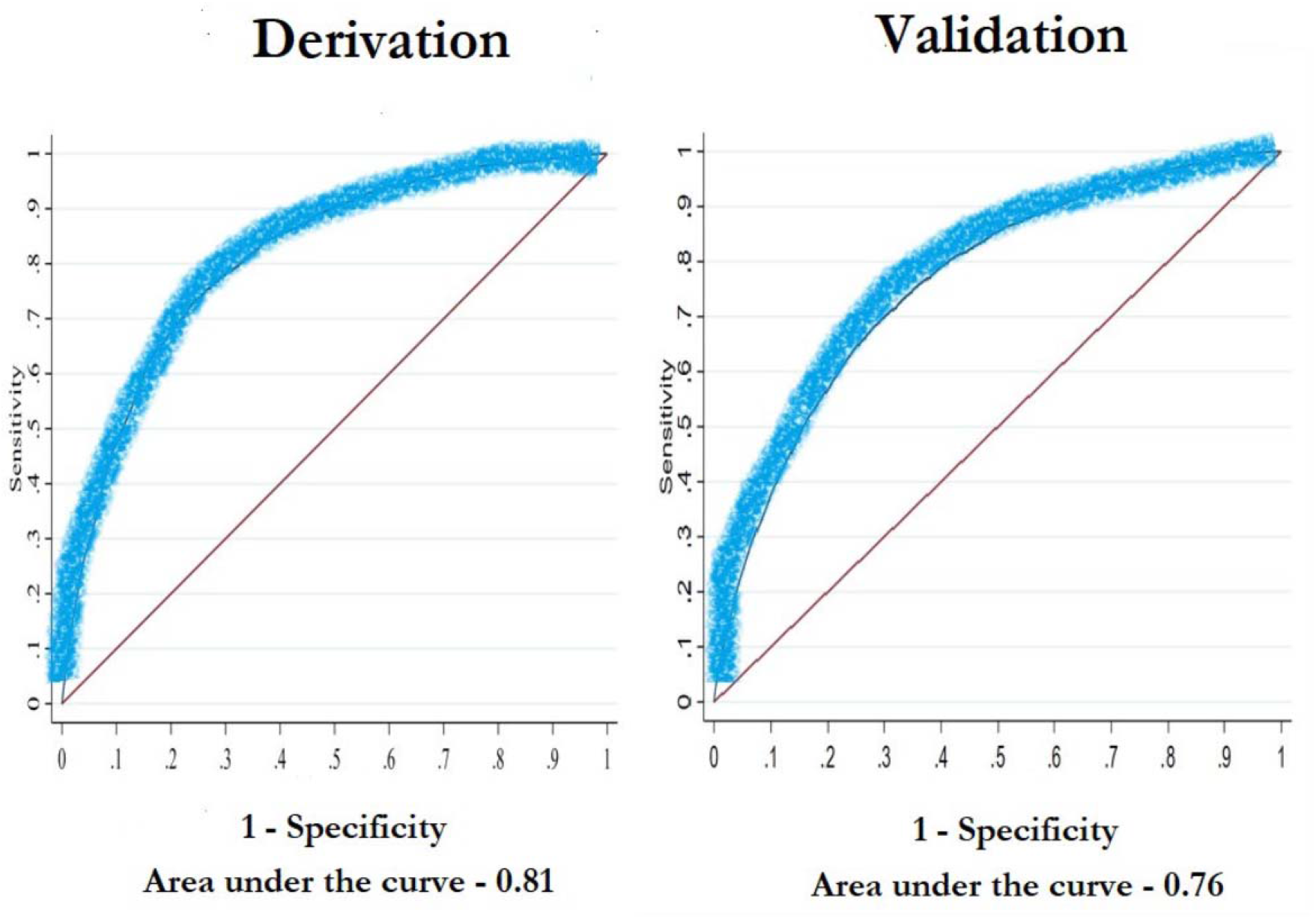
Time-dependent ROC curves predicting 1-year death adjusted for the linear predictor with the SWEDHEART registry (Derivation dataset) & the Western Denmark registry (Validation dataset)

**Figure 2.**
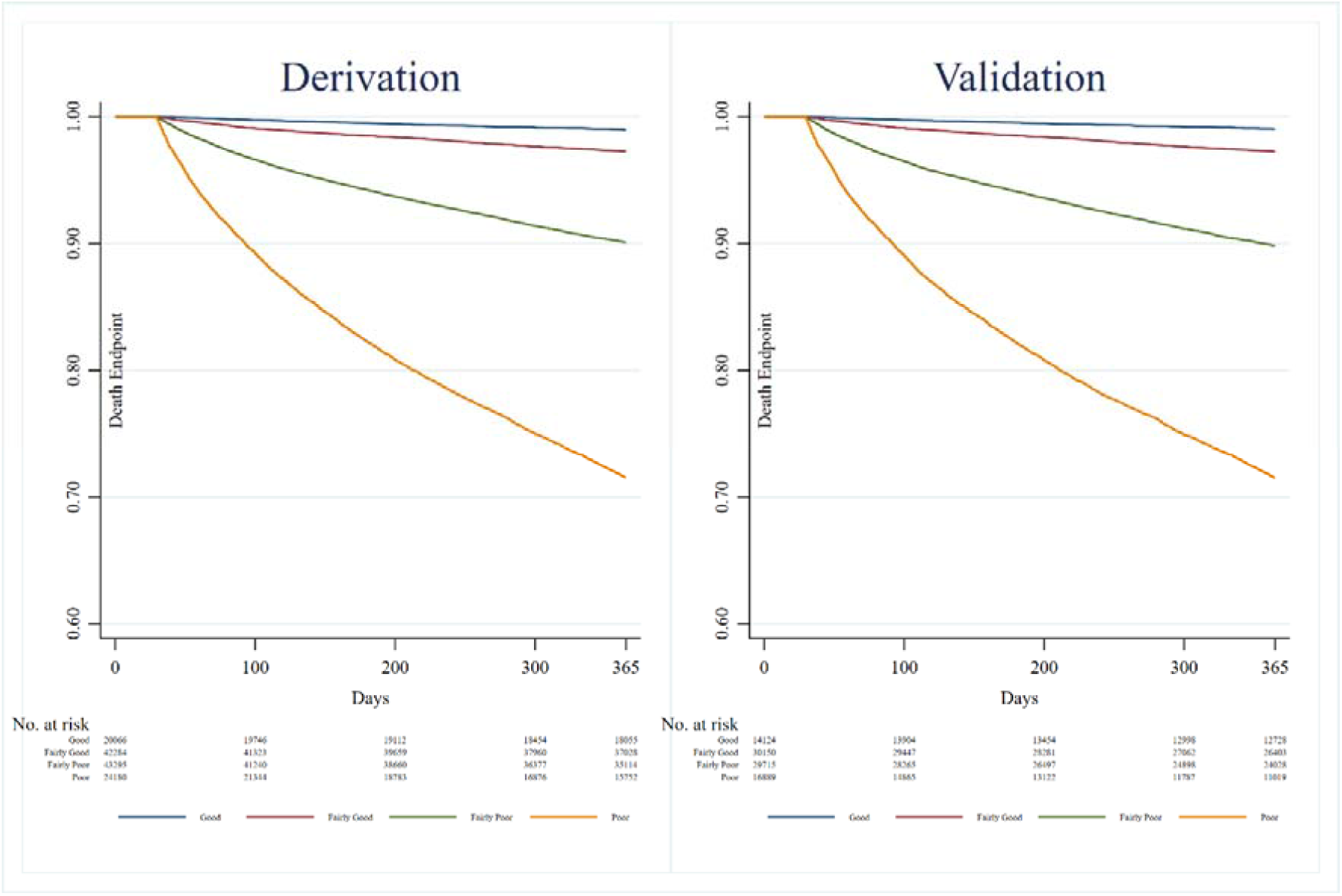
Kaplan-Meier curves of the categorized linear predictor (weighted sum of regression coefficients produced from the adjusted Cox model divided into groups) from SWEDEHEART registry (Derivation dataset) & Western Denmark registry (Validation dataset)

**Figure 3.** Histograms of the categorized linear predictor from the SWEDEHEART registry (Derivation dataset) & Western Denmark registry (Validation dataset) Appendix

**Figure 4.**
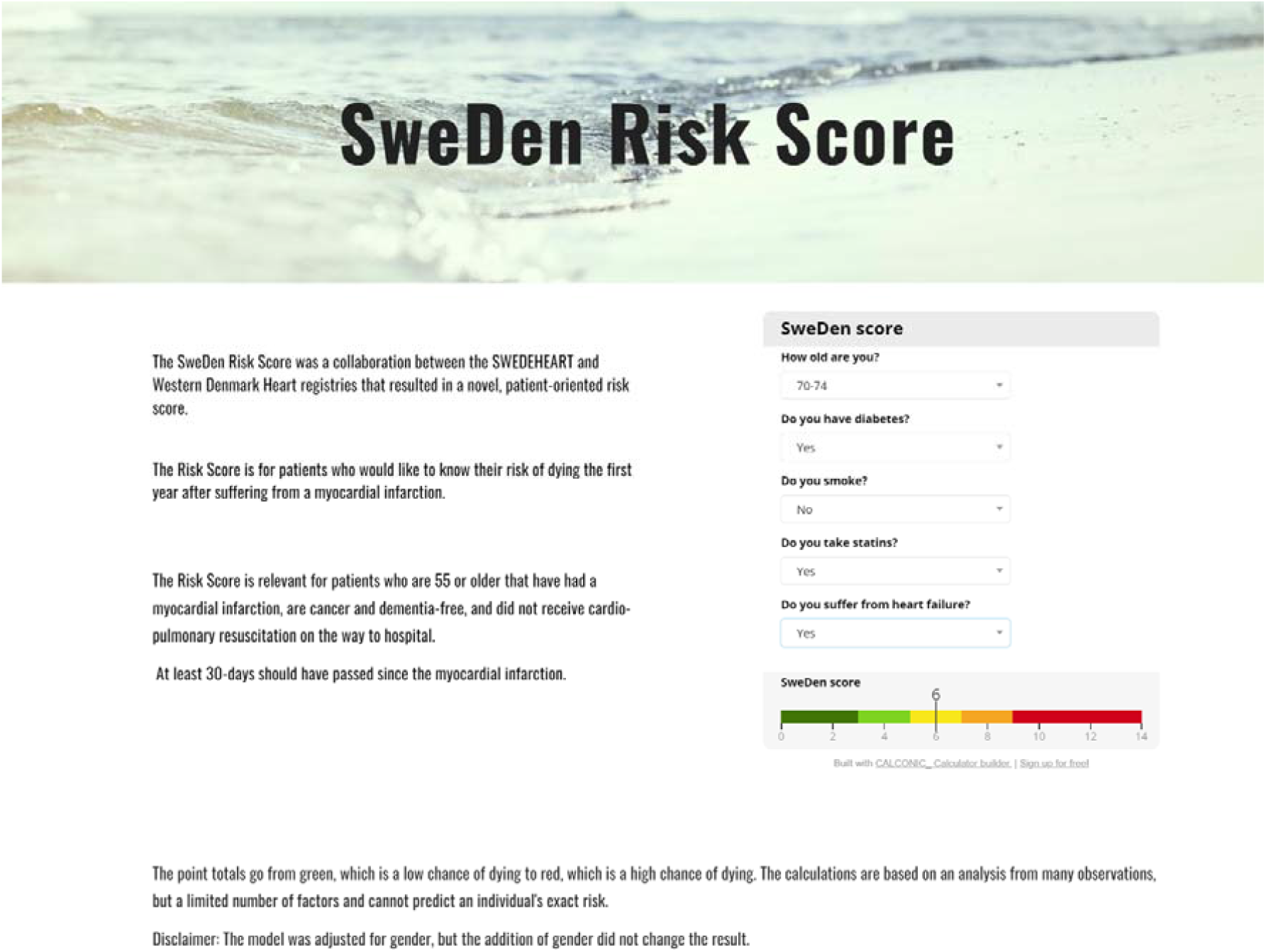
SweDen risk score calculator

## Discussion

The SweDen score is a patient-oriented risk score with an AUC of 0.81 in the derivation cohort and 0.76 in the validation cohort. Despite the simplicity of the SweDen score, the AUC was high, the estimates were reproducible in a different cohort, and the results suggested both good discrimination and calibration.

The TIMI and GRACE scores are two other, in this context, meaningful scores that had the same aim as the SweDen score. The C-statistic from the GRACE score for 1-year mortality was 0.82 (95% CI 0.79-0.84)^10^ and TIMI score was 0.65 (95% CI 0.63-0.66)^3^ making the SweDen risk score a viable alternative for patients themselves to use. The chosen factors in these different scores are debatable. The SweDen risk score incorporated diabetes and previous and current smokers as separate factors, while the TIMI score only includes diabetes and current smokers if these are part of a combination of at least 3 factors. ^11 12^

While systolic blood pressure was a factor in the GRACE score, we chose not to include it in the SweDen score because daily fluctuations in blood pressure would need to be accounted for rather than selecting one random daily measurement.^13^ Killip class was used in the GRACE score as well, which may have increased the prediction accuracy in the SweDen score^14^ if included, but it is a value unknown to most patients. Furthermore, if more predictors would have been included from the SWEDEHEART registry to predict death 1-year following a MI, prediction accuracy may have increased. Other SWEDEHEART studies have demonstrated this applying machine learning algorithms.^15^ However, the calibration slope in the validation dataset was 1.03 indicating sufficiently high prediction accuracy.

Prediction accuracy via the calibration slope as well as the harmonious estimates show that the external validation was successful. Unfortunately, not enough studies engage in the transportability of a risk equation to a new population in cardiovascular disease.^16^ External validation is crucial to evaluate a model’s reproducibility and that is why the SweDen risk score was validated externally with the Western Denmark Heart Registry.^5^

In summary, we wanted to create a patient-oriented risk score that predicts the risk of death within 1-year after suffering a MI. This was developed in collaboration between Sweden and Denmark resulting in the validated patient-oriented SweDen risk score. The SweDen risk score includes less factors than other similar risk scores, but has a predictability that we found to be at as good as other risk scores recommended in current guidelines. A further advantage is that patients themselves can fill in their information and visualize the potential benefit of smoking cessation and statin use, making it a feasible tool for patients who have suffered a MI.

## Supporting information

Figure 3, Table 3, Calculation of risk score

## Data Availability

Researchers interested in further analyses of the data can contact the authors.

## Notes

### Competing Interest Statement

The authors have declared no competing interest.

### Funding Statement

This study was funded by the Swedish Research council and the Swedish HeartLung foundation.

### Author Declarations

The study was approved by the ethics committee in Lund, 2015, 297-approval code. The data is from the SWEDEHEART registry and individuals are automatically included in registries unless they opt out.

