## Supplementary material for "The SweDen risk score- predicting death 1-year after myocardial infarction": Figure 3, Table 3, Calculation of risk score

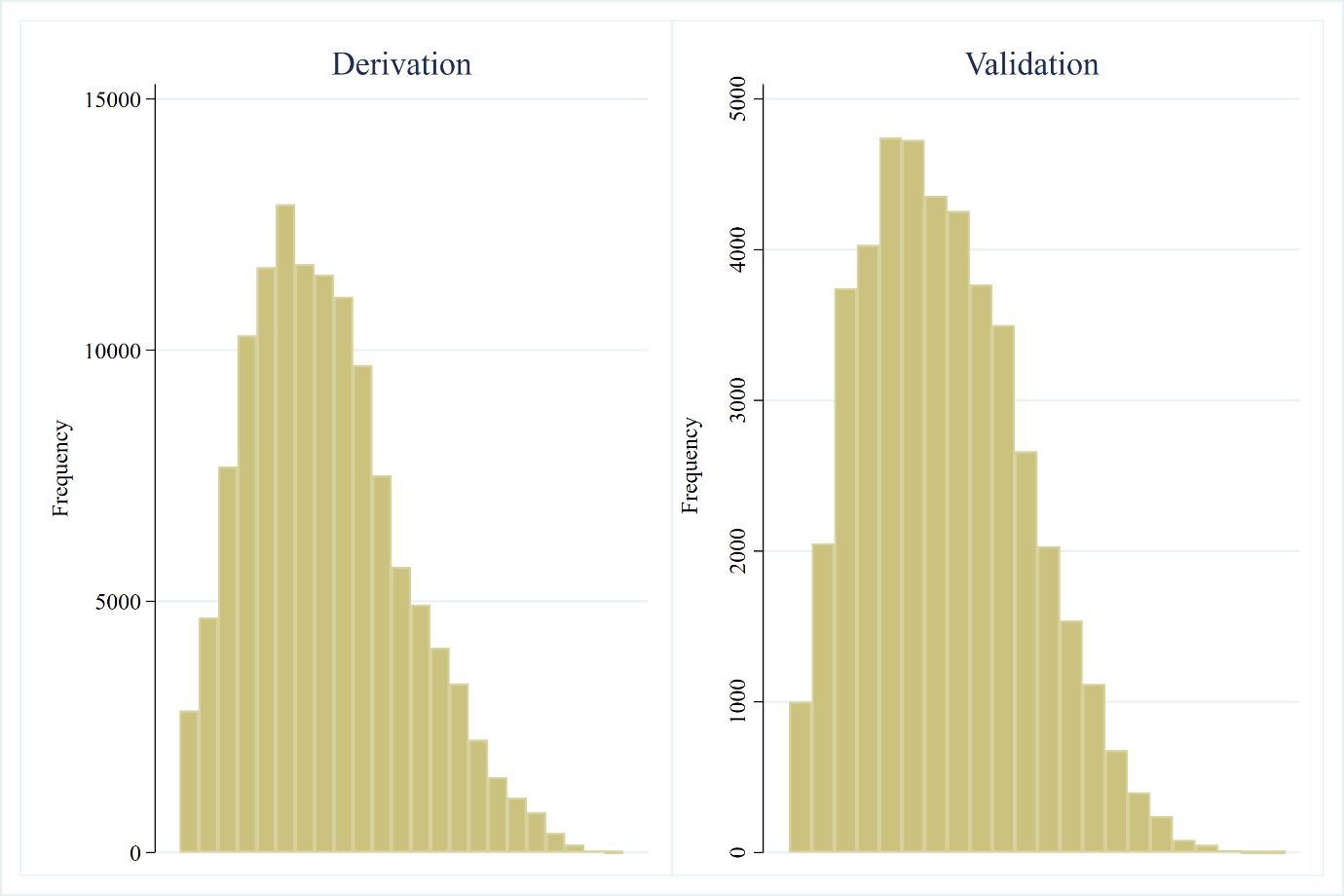

Figure 3. Histograms of the linear predictor

Table 3.

| Measure | Derivation data | | Validation data | |
| --- | --- | --- | --- | --- |
|  | Estimate | se | Estimate | Se |
| HR; Fairly good vs. good* | 2.78 | .217 | 2.55 | .376 |
| HR; Fairly bad vs. good* | 10.45 | .772 | 7.47 | 1.047 |
| HR; Bad vs. good* | 33.69 | 2.466 | 21.27 | 2.965 |

HR; hazard ratios

Calculation of risk score

Step 1. Cox proportional hazards regression log hazard coefficients

Risk factor	Coefficient, B*_i_*	Mean or proportion

Age, years	.07801	73.3

Gender	.08949	.64

Diabetes	.51539	.22

Smoker previous	.03841	.35

Smoker current	.38103	.17

Smoker non	Base	.48

Statins not prescribed	.65660	.13

Heart failure	.96994	.14

Step 2. Definition of a point

One point constitutes:

5 years aging multiplied by the coefficient for Age

From our model, we have *B_i_* = .07801*5= 0.39005

3. Formula for points

Categories	Reference value(*W_ij_*)	*B_i_*	Points*ij*=*B_i_(W_ij_-W_i_REF)/B*

.07801

Age	55-59	57= *W_1_REF*	0

60-64	62	1

65-69	67	2

70-74	72	3

75-79	77	4

80-84	82	5

85-89	87	6

90-94	92	7

95-105	97	8

Gender	Female	0=W_2_REF	.08949	0

Male	1	0

Diabetes	No	0=W_3_REF	.51539	0

Yes	1	1

Smoker previous	No	0=W_4_REF	.03841	0

Yes	1	0

Smoker current	No	0=W_4_REF	.38103	0

Yes	1	1

Statins	Prescribed	0=W_5_REF	.65660	0

Not prescribed	1	2

Heart failure	No	0=W_6_REF	.96994	0

Yes	1	2

4. Point System

Risk factor	Categories	Points

Age	55-59	0

60-64	1

65-69	2

70-74	3

75-79	4

80-84	5

85-89	6

90-94	7

95-105	8

Gender	Female	0

Male	0

Diabetes	No	0

Yes	1

Smoker previous	No	0

Yes	0

Smoker current	No	0

Yes	1

Statins	Prescribed	0

Not prescribed	2

Heart failure	No	0

Yes	2

5. Attach risk to every point

$\sum_{i=1}^{p} B_{i}\overline{X}_{i}$ = .0780096*(73.3) + .0894894*(0.64) + .5153974*(0.23) + .0384073*(0.35) + .3810368*(0.17) +.6566036*(0.13) + .9699417*(0.14) = 6.1932874

$\sum_{i=1}^{p} B_{i}X_{i}$ ≈ for every point

.0780096*(57)+.390048*(0) = 4.4465472

.0780096*(57)+.390048*(1) = 4.8365952

.0780096*(57)+.390048*(2) = 5.2266432

.0780096*(57)+.390048*(3) = 5.6166912

.0780096*(57)+.390048*(4) = 6.0067392

.0780096*(57)+.390048*(5) = 6.3967872

.0780096*(57)+.390048*(6) = 6.7868352

.0780096*(57)+.390048*(7) = 7.1768832

.0780096*(57)+.390048*(8) = 7.5669312

.0780096*(57)+.390048*(9) = 7.9569792

.0780096*(57)+.390048*(10) = 8.3470272

.0780096*(57)+.390048*(11) = 8.7370752

.0780096*(57)+.390048*(12) = 9.1271232

.0780096*(57)+.390048*(13) = 9.5171712

.0780096*(57)+.390048*(14) = 9.9072192

$S_{o}(t)$= 0.9035 Survival at one-year

5) Attach risk estimate to each point total risk=$1-S_{o}\left( t \right)^{{exp}^{\left( \sum_{i=1}^{p} B_{i}X_{i}-\sum_{i=1}^{p} B_{i}\overline{X}_{i} \right)}}$

Points	Risks associated with point totals

0	0.0176

1	0.0259

2	0.0380

3	0.0557

4	0.0812

5	0.1176

6	0.1687

7	0.2389

8	0.3318

9	0.4488

10	0.5851

11	0.7273

12	0.8533

13	0.9412

14	0.9848
